# Evaluating differential item functioning in the EQ-5D-5L in acute ischemic stroke

**DOI:** 10.64898/2026.03.10.26348094

**Authors:** Olayinka I. Arimoro, Ayoola Ademola, Michael D. Hill, Bijoy K. Menon, Tolulope T. Sajobi

**Affiliations:** Department of Community Health Sciences & O’Brien Institute for Public Health, Cumming School of Medicine, University of Calgary, Calgary, AB, Canada; Cancer Analytics Division, Alberta Health Services, Calgary, AB, Canada; Department of Surgery, Cumming School of Medicine, University of Calgary, Calgary, AB, Canada; Department of Clinical Neurosciences & Hotchkiss Brain Institute, Cumming School of Medicine, University of Calgary, Calgary, AB, Canada

**Keywords:** differential item functioning, EQ-5D-5L, health-related quality of life, graded response model, stroke, patient-reported outcomes, measurement invariance, item response theory

## Abstract

**Background:** Health-related quality of life is a key secondary endpoint in stroke trials. Differential item functioning (DIF) occurs when individuals with the same underlying HRQOL interpret and respond differently to questionnaire items due to group characteristics, potentially biasing treatment comparisons. This study evaluates DIF in the patient-reported five-level EuroQOL (EQ-5D-5L) among patients with acute ischemic stroke across age, sex, and treatment groups.

**Methods:** Data were obtained from the AcT trial, a registry-based randomized comparison of alteplase and tenecteplase. Patients completed the EQ-5D-5L at 90 days post-stroke. DIF was assessed using multigroup graded response models with the Wald-based sweep procedure, which accounts for between-group differences in latent trait distributions. We quantified effect sizes using signed weighted area between curves (sWABC), considering |sWABC| <0.10 as negligible.

**Results:** Among 1,264 patients (51.2% tenecteplase; 46.5% female; 30.1% aged ≥80). Omnibus testing revealed significant DIF only for age (*X*² = 86.9, *p* < 0.001); neither sex (*X*² = 31.7, *p* = 0.063) nor treatment (*X*² = 22.4, *p* = 0.379) showed evidence of DIF. At the item level, four items flagged for age-related DIF: self-care, usual activities, pain/discomfort, and anxiety/depression. However, only self-care (sWABC = -0.46) and usual activities (sWABC = - 0.34) showed moderate effects, while pain/discomfort (sWABC = -0.002) and anxiety/depression (sWABC = 0.09) were negligible. Importantly, factor scores from models with and without DIF adjustment correlated (correlation coefficient = 0.98).

**Conclusions:** The EQ-5D-5L appears to function equivalently across sex and treatment groups in this stroke population. Age-related DIF, though statistically detectable in physical functioning items, had little practical consequence for individual scores, findings that support the instrument’s use for HRQOL comparisons in stroke trials.

**Registration:** URL: https://www.clinicaltrials.gov; Unique identifier: NCT03889249.

## Introduction

Stroke is a leading cause of death and disability.^1,2^ According to the World Health Organization, approximately 15 million individuals experience stroke globally each year.^3^ Of these, 5 million will die, and another 5 million will be left with permanent disability, placing a substantial burden on families and caregivers.^3^ Ischemic stroke occurs when blood flow to the brain is interrupted, causing mild to severe impairments in physical, cognitive, and emotional functioning.^4,5^ These impairments affect multiple domains of health-related quality of life (HRQOL), including cognition, physical functioning, pain, anxiety, fatigue, and depression^6^. These HRQOL domains have been shown to be patient-reported outcomes that matter most to individuals who have survived stroke.^6^ As stroke incidence continues to rise globally, assessing these outcomes has become essential for optimizing recovery and rehabilitation.

Patient-reported outcome measures (PROMs) provide direct insight into how patients perceive their health and quality of life.^7,8^ Regulatory bodies, including the United States Food and Drug Administration^9^ and the European Medicines Agency,^10^ have promoted PROMs as clinical endpoints in randomized controlled trials.^9–11^ In addition to demonstrating therapeutic benefits on conventional clinical outcomes, regulatory agencies now require evidence of treatment efficacy on patient-reported health status.^12^ In acute stroke trials reporting PROMs as secondary endpoints, the five-level EuroQOL questionnaire (EQ-5D-5L)^13^ is the most widely adopted measure.^14,15^

Although psychometric properties such as validity and reliability guide PROM selection in clinical trials, measures used in stroke trials are rarely evaluated for differential item functioning (DIF).^16,17^ DIF occurs when individuals with the same underlying HRQOL interpret and respond to questionnaire items differently based on patient characteristics such as age or sex.^17^ When DIF is present, comparisons of HRQOL across patient subgroups may be confounded by measurement artifacts rather than true health differences, leading to biased treatment comparisons, reduced statistical power, and erroneous conclusions about treatment effects.^18,19^

Heterogeneity in HRQOL assessments due to age and sex differences has been documented in acute stroke trials.^20–23^ Female stroke survivors report lower HRQOL than males, regardless of stroke severity or treatment received,^20–22^ while older patients report poorer HRQOL than younger patients.^20,23^ However, the extent to which these disparities reflect true health differences versus DIF in the measurement instrument remains unclear. While DIF in the EQ-5D has been investigated previously,^24–26^ most analyses were conducted in observational studies and did not explore DIF’s potential impact on treatment effect conclusions in randomized controlled trials. This study evaluated the presence and magnitude of DIF in EQ-5D-5L items in an acute ischemic stroke trial across sex, age, and treatment subgroups.

## Methods

### Data Source

Data were from the AcT trial (Alteplase Compared to Tenecteplase), a pragmatic, multicenter, parallel-group, open-label with blinded endpoint assessment (PROBE), registry-linked randomized trial evaluating the non-inferiority of intravenous tenecteplase compared with alteplase in patients with acute ischemic stroke (ClinicalTrials.gov NCT03889249).^27,28^ Patients were eligible if they were 18 years or older with ischemic stroke causing disabling neurological deficit, presenting within 4.5 hours of symptom onset, and eligible for thrombolysis per Canadian Stroke Best Practice Recommendations.^29^ The trial enrolled 1,600 patients from 22 primary and comprehensive stroke centers across Canada between December 2019 and January 2022; 1,577 were included in the intention-to-treat analysis. Two centers in Quebec, Canada, used prospective written or verbal consent from patients or their representatives. Patients or their legal representatives at remaining centres provided written or electronic informed consent as soon as possible after treatment, within seven days of randomization, or before discharge, whichever was earlier.HRQOL at 90 days was assessed by telephone interview using the EQ-5D-5L.^27^ Although cognitive impairment was not formally assessed, patients with severe pre-morbid disability were excluded. Additional details have been published previously.^27,28^ The trial was regulated by Health Canada (Clinical Trials Application number 231509) and by research ethics boards at participating centres.

### EQ-5D-5L Measure

The EQ-5D-5L is a standardized generic instrument for measuring HRQOL, introduced by the EuroQol Group in 2009 to improve sensitivity and reduce ceiling effects compared to the three-level version.^30,31^ It consists of five dimensions: mobility, self-care, usual activities, pain/discomfort, and anxiety/depression, each with five response levels ranging from 1 (no problems) to 5 (extreme problems/unable to perform).^30^

### Statistical Analysis

Analyses included patients with complete EQ-5D-5L responses at 90 days. Baseline characteristics were summarized using medians with interquartile ranges (IQR) for continuous variables and frequencies with percentages for categorical variables. Sparse response categories (<2.5% endorsement) were merged with adjacent categories to ensure stable parameter estimation.

We used a graded response model (GRM),^32^ an item response theory approach for ordinal data, to evaluate whether EQ-5D-5L items function equivalently across patient subgroups. Model adequacy was assessed using standard fit indices^33,34^: comparative fit index (CFI ≥0.95 excellent, ≥0.90 acceptable), Tucker-Lewis index (TLI ≥0.95 excellent, ≥0.90 acceptable), and standardized root mean square residual (SRMSR ≤0.05 excellent, ≤0.08 acceptable). We note that root mean square error of approximation (RMSEA) derived from limited-information fit statistics tends to be inflated for instruments with few items and many response categories;^35^ therefore, we prioritized CFI, TLI, and SRMSR for fit evaluation. Local dependence between item pairs was assessed using Chen and Thissen’s G² statistic with Benjamini-Hochberg correction;^36,37^ standardized residual correlations exceeding |0.20| would indicate meaningful violations.

DIF was evaluated using multigroup models comparing item responses across age (<80 vs ≥80 years), sex (male vs female), and treatment (alteplase vs tenecteplase) groups. The age threshold of 80 years was selected for consistency with prior AcT trial analyses.^20,27^ The Wald-based sweep procedure with structural parameter adjustment was used, which accounts for true differences in underlying health status between groups when testing for item bias.^38,39^ This two-stage approach first identifies anchor items (those functioning equivalently across groups), then tests remaining items while controlling for group differences in latent trait distributions^38^.

Effect sizes were quantified using the signed weighted area between curves (sWABC), which measures the practical magnitude of item-level DIF.^39,40^ Following established guidelines, |sWABC| <0.10 was considered negligible, 0.10–0.29 small, 0.30–0.49 moderate, and ≥0.50 large.^39^ To assess practical impact on individual patients, we compared factor scores from models with and without DIF adjustment; a high correlation would indicate minimal impact on individual-level assessment. To complement item-level analyses, differential test functioning (DTF) was calculated, which assesses measurement bias at the scale level.^41,42^ Signed DTF (sDTF) represents the expected score difference between groups at the same latent HRQOL level, while unsigned DTF (uDTF) captures total bias magnitude regardless of direction.

All analyses were performed using R version 4.5.1^43^ with the *mirt* package.^44^

## Results

Of 1,577 patients in the AcT trial, 241 (15.3%) died before 90-day follow-up. Among the 1,264 patients with complete EQ-5D-5L data, 647 (51.2%) received tenecteplase, 588 (46.5%) were female, and 380 (30.1%) were aged ≥80 years (Table 1). The median age was 72.0 years (IQR: 61.0–81.0). Response distributions for each EQ-5D-5L item are presented in Supplementary Table S1. Categories 4 and 5 were collapsed for pain/discomfort and anxiety/depression due to sparse endorsement of extreme responses. Response category 5 of pain/discomfort and anxiety/depression were 6 (0.5%) and 7 (0.6%), respectively.

**Table 1.** Descriptive characteristics of the study cohort (N = 1,264)

| Patient Characteristics | n (%) |
| --- | --- |
| Sex |  |
| *Female* | 588 (46.5) |
| *Male* | 676 (53.5) |
| Type of registry |  |
| *OPTIMISE* | 703 (55.6) |
| *QUICR* | 561 (44.4) |
| Type of enrolling centre |  |
| *Primary stroke centre* | 86 (6.8) |
| *Comprehensive stroke centre* | 1,178 (93.2) |
| Treatment received |  |
| *Tenecteplase (TNKase™, Metalyse™)* | 647 (51.2) |
| *Alteplase (Activase™)* | 617 (48.8) |
| Endovascular thrombectomy (EVT) |  |
| *No* | 860 (68.0) |
| *Yes* | 404 (32.0) |
| mRS 0-1 at 90 days |  |
| *No* | 718 (56.8) |
| *Yes* | 546 (43.2) |
| Return to pre-stroke status |  |
| *No* | 860 (68.0) |
| *Yes* | 404 (32.0) |
|  | **Median (IQR)** |
| Age, years | 72.0 (61.0–81.0) |
| NIHSS<sup>§</sup> | 9.0 (5.0–15.0) |
| Onset to needle time, minutes<sup>§</sup> | 128.5 (93.0–185.0) |

**Note:** IQR = interquartile range; mRS = modified Rankin Scale; Stroke severity was measured using the National Institutes of Health Stroke Scale (NIHSS). <sup>§</sup>Indicates patient characteristics with missing values: NIHSS = 4 (0.3%); Onset to needle time = 10 (0.8%).

The GRM demonstrated acceptable fit (CFI = 0.97, TLI = 0.93, SRMSR = 0.07). Item discrimination parameters ranged from 0.99 to 6.38, with physical functioning items (mobility, self-care, usual activities) showing stronger relationships with underlying HRQOL than symptom items (pain/discomfort, anxiety/depression) (Table 2). Four item pairs showed statistically significant local dependence after correction for multiple testing: mobility–self-care, mobility–usual activities, usual activities–pain/discomfort, and pain/discomfort–anxiety/depression (see Supplementary Table S2). However, standardized residual correlations were uniformly small (range: -0.12 to 0.12), well below the |0.20| threshold indicating meaningful dependence (see Supplementary Figure S1). This pattern reflects expected overlap among physical functioning items and between symptom items, without threatening the validity of subsequent analyses.

**Table 2.**
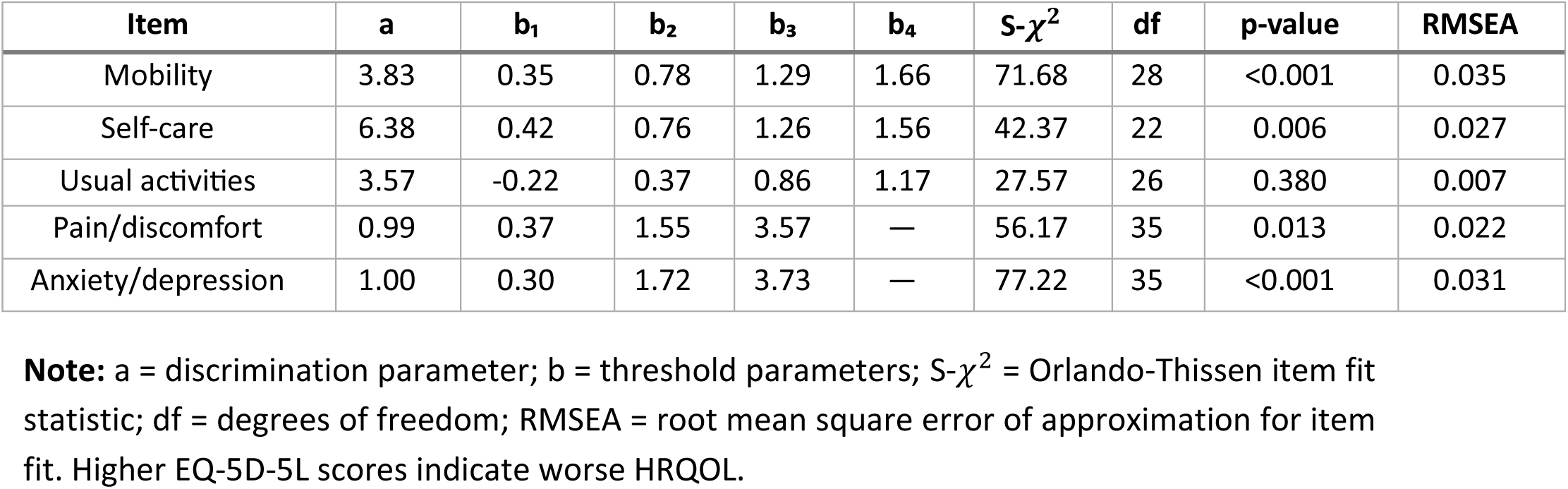
Item parameters and fit statistics for the graded response model.

Table 3 presents omnibus DIF test results. Significant DIF was detected for age (*X*² = 86.9, df = 21, *p* < 0.001), but not for sex (*X*² = 31.7, df = 21, *p* = 0.063) or treatment (*X*² = 22.4, df = 21, p = 0.379). Item-level results for sex and treatment are provided in Supplementary Table S3. Item-level analysis for age identified four items with statistically significant DIF after Benjamini-Hochberg correction: self-care, usual activities, pain/discomfort, and anxiety/depression (Table 4). Mobility did not show significant DIF (adjusted p = 0.256). However, statistical significance diverged from practical significance. Effect sizes were moderate for self-care (sWABC = -0.46) and usual activities (sWABC = -0.34), indicating that older adults (≥80 years) reported greater difficulty on these items than younger adults with equivalent underlying HRQOL. In contrast, pain/discomfort and anxiety/depression showed negligible effect sizes (sWABC = -0.002 and 0.09, respectively), meaning the statistically detected DIF would have no practical impact.

**Table 3.** Differential item functioning analysis results: Omnibus DIF tests by grouping variable.

| Grouping Variable | Configural AIC | Constrained AIC | ΔAIC | $\chi^2$ | df | p-value | Conclusion |
| --- | --- | --- | --- | --- | --- | --- | --- |
| Age (<80 vs ≥80 years) | 12,432.2 | 12,477.1 | -44.9 | 86.91 | 21 | <0.001 | Significant DIF |
| Sex (Male vs Female) | 12,494.4 | 12,484.1 | +10.3 | 31.71 | 21 | 0.063 | No significant DIF |
| Treatment (tPA vs TNK) | 12,522.6 | 12,503.0 | +19.6 | 22.36 | 21 | 0.379 | No significant DIF |

**Note:** DIF = differential item functioning; df = degrees of freedom; AIC = Akaike Information Criterion; $\Delta$ AIC = Configural AIC minus Constrained AIC (negative values favor the configural model, indicating DIF). Omnibus test compares configural model (free item parameters across groups) to constrained model (equal item parameters) using the likelihood ratio test. Significance level $\alpha = 0.05$ .

**Table 4.** Item-level differential item functioning results for age (<80 vs ≥80 years)

| Item | $\chi^2$ | df | p-value | Adj. p | ΔAIC | ΔBIC | sWABC | Effect Size |
| --- | --- | --- | --- | --- | --- | --- | --- | --- |
| Mobility | 5.32 | 4 | 0.256 | 0.256 | 2.68 | 23.25 | -0.35 | Moderate |
| Self-care | 27.74 | 4 | <0.001 | <0.001 | -19.74 | 0.83 | -0.46 | Moderate |
| Usual activities | 15.10 | 4 | 0.004 | 0.006 | -7.10 | 13.47 | -0.34 | Moderate |
| Pain/discomfort | 16.21 | 4 | 0.003 | 0.005 | -8.21 | 12.36 | -0.002 | Negligible |
| Anxiety/depression | 34.75 | 4 | <0.001 | <0.001 | -26.75 | -6.18 | 0.09 | Negligible |

**Note:** DIF = differential item functioning; $\chi^2$ = chi-square statistic; df = degrees of freedom; Adj. p = Benjamini-Hochberg adjusted p-value; AIC = Akaike Information Criterion; BIC = Bayesian Information Criterion; $\Delta$ AIC and $\Delta$ BIC = difference in information criteria when freeing item parameters (negative values indicate improved fit with DIF); sWABC = signed weighted area between curves. Effect size interpretation: |sWABC| <0.10 = negligible, 0.10–0.29 = small, 0.30–0.49 = moderate, ≥0.50 = large. Negative sWABC indicates older adults (≥80 years) report greater difficulty than younger adults at the same underlying HRQOL level. Higher EQ-5D-5L scores indicate worse health status.

Factor scores from DIF-adjusted and unadjusted models were highly correlated (correlation coefficient = 0.98), indicating that age-related DIF had a modest impact on individual-level HRQOL assessment (Figure 1). Score differences were concentrated among older patients (≥80 years: mean difference = 0.37, SD = 0.06), while younger patients showed essentially no impact (<80 years: mean difference ≈ -0.0001, SD = 0.02). Positive differences indicate that the unadjusted model slightly overestimated HRQOL impairment in older patients compared to the DIF-adjusted model. Despite this systematic pattern, the high correlation between score sets suggests minimal practical consequence for clinical interpretation.

**Figure 1.**
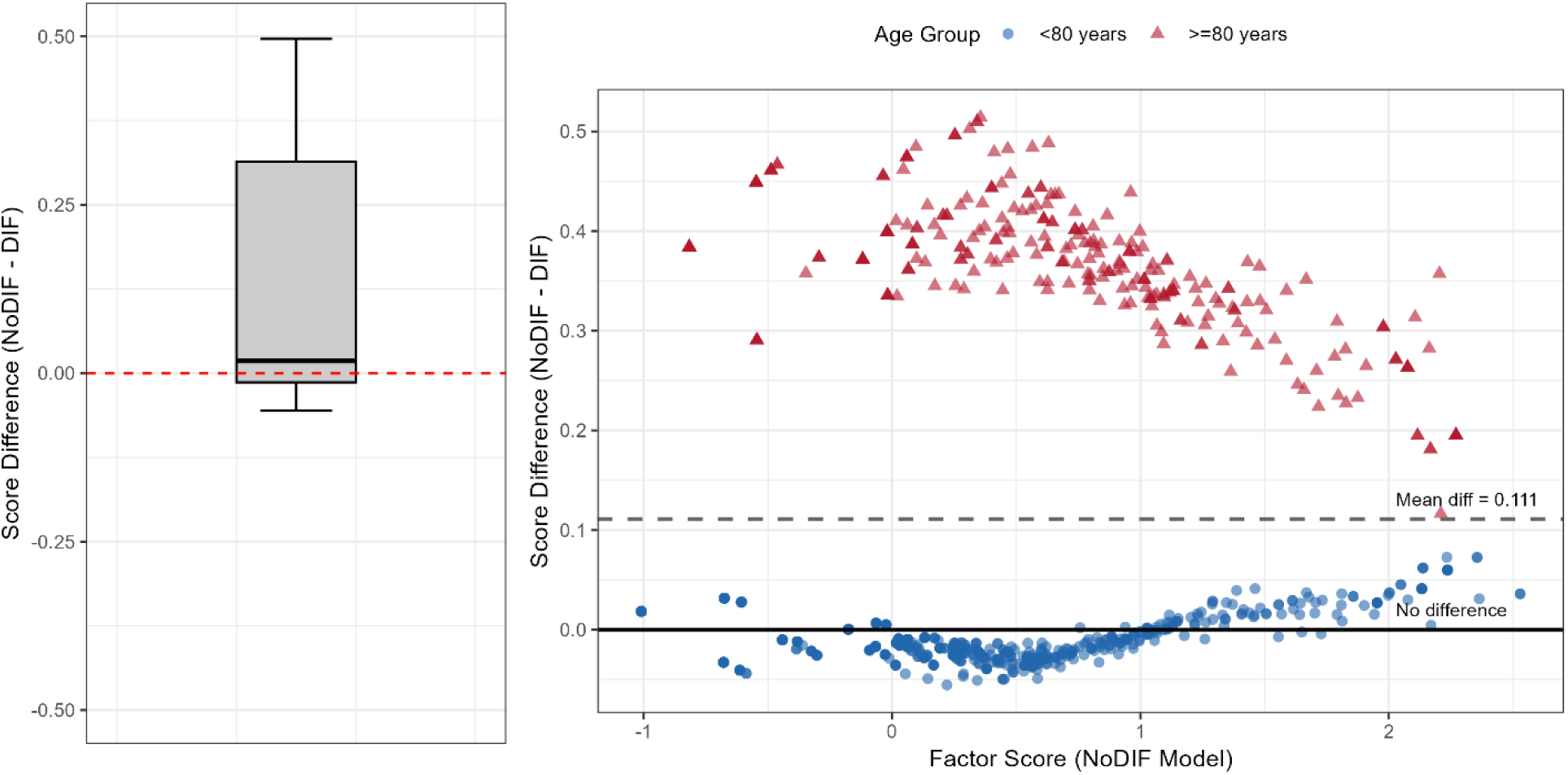
Impact of age-related differential item functioning on factor scores **Note:** NoDIF = model assuming measurement invariance (equal item parameters across age groups); DIF = model allowing item parameters to differ between age groups (<80 years vs ≥80 years). Left panel: Boxplot of factor score differences (NoDIF − DIF). The median difference was 0.02 (IQR: -0.17 to 0.18), with most differences clustered near zero. Right panel: Scatterplot of score differences against NoDIF factor scores, stratified by age group. The solid horizontal line indicates no difference (0.00); the dashed line indicates the mean difference (0.11). Younger patients (<80 years; blue circles) show minimal score differences clustered tightly around zero across the range of factor scores. Older patients (≥80 years; red triangles) demonstrate systematically positive differences, indicating the NoDIF model yielded higher (worse) HRQOL scores compared to the DIF-adjusted model. This pattern suggests that ignoring age-related DIF slightly overestimates HRQOL impairment in older stroke patients. Despite this systematic pattern, the high correlation between score sets (correlation coefficient = 0.98) indicates the practical impact on individual-level assessment is modest. Higher EQ-5D-5L factor scores indicate worse health-related quality of life.

DTF analysis revealed a signed DTF of -1.06 (5.3% of the 0–20 scale range), indicating that at the same underlying HRQOL level, older patients (≥80 years) would score approximately 1 point higher (worse) on the summed EQ-5D-5L than younger patients. The unsigned DTF was 1.24, suggesting that DIF effects were predominantly in the same direction (favoring younger patients) rather than canceling across items. These scale-level findings corroborate the item-level results; while statistically detectable, the overall measurement bias attributable to age-related DIF is modest.

## Discussion

In this study of 1,264 acute ischemic stroke patients from the AcT trial, we found that the EQ-5D-5L demonstrates reasonable measurement invariance across age, sex, and treatment groups. While statistically significant age-related DIF was detected in four items, the practical impact on individual HRQOL scores was modest, as evidenced by the high correlation (correlation coefficient = 0.98) between DIF-adjusted and unadjusted factor scores. No meaningful DIF was observed across sex or treatment groups, supporting the validity of EQ-5D-5L comparisons in stroke trials.

These findings have direct relevance for stroke clinical practice and research. The EQ-5D-5L is increasingly used not only as a secondary endpoint in stroke trials but also for computing quality-adjusted life years in health technology assessments and cost-effectiveness analyses.^15,45^ Our results suggest that observed differences in EQ-5D-5L scores between patient subgroups, whether by age, sex, or treatment, more likely reflect genuine differences in health status rather than measurement artifact. This supports the continued use of the EQ-5D-5L for HRQOL assessment in heterogeneous stroke populations without requiring group-specific scoring adjustments.

The absence of treatment-related DIF is particularly reassuring for the AcT trial’s secondary HRQOL findings. Patients receiving tenecteplase and alteplase interpreted and responded to EQ-5D-5L items equivalently, meaning any observed differences in HRQOL between treatment arms can be attributed to true treatment effects rather than differential item functioning. The pattern of age-related DIF warrants careful interpretation. Four items showed statistically significant DIF, yet effect sizes varied considerably. Self-care and usual activities demonstrated moderate effects (sWABC = -0.46 and -0.34), indicating older patients reported greater difficulty on these items than younger patients with equivalent underlying HRQOL. However, pain/discomfort and anxiety/depression showed negligible effects despite statistical significance. This divergence highlights the importance of quantifying DIF magnitude rather than relying solely on statistical tests, particularly in large samples where trivial effects can achieve significance.^46,47^

Notably, the items showing meaningful age-related DIF (self-care and usual activities) assess physical functioning domains where age-related expectations may influence patient responses. An 80-year-old patient may interpret *“some problems washing or dressing“* differently than a 60-year-old, not because the item is biased, but because baseline expectations differ. Whether this represents true DIF or appropriate adaptation to age-related functional norms remains debatable. Regardless, the minimal impact on overall scores (mean difference = 0.37 for older patients, essentially zero for younger patients) suggests this does not threaten the validity of age-stratified comparisons in clinical trials.

Our findings align with previous psychometric studies of the EQ-5D in stroke populations. Whynes et al. examined DIF in the EQ-5D using data from a large stroke trial and found no significant DIF between patient and proxy reports, though regional differences were observed.^25^ Golicki et al. demonstrated the validity of the EQ-5D-5L in Polish stroke patients,^48^ and Chen et al. confirmed its responsiveness in Taiwanese stroke survivors undergoing rehabilitation.^49^ These studies support the instrument’s measurement properties across diverse stroke populations.

This study has several methodological strengths. We employed the Wald-based DIF sweep procedure with structural parameter adjustment, which accounts for true differences in underlying health status between groups when testing for item bias. This approach is a critical refinement over simpler approaches that may confound group means differences with item- level bias. Local dependence was assessed using Chen and Thissen’s G² statistic with Benjamini-Hochberg correction, and effect sizes were quantified using signed weighted area between curves rather than statistics designed for continuous outcomes.

Several limitations should be acknowledged. First, the EQ-5D-5L contains only five items, which constrains DIF detection methodology. With few items, anchor identification becomes challenging, and the power to distinguish true DIF from random variation is limited. As Zumbo noted,^50^ DIF analyses with very short instruments face inherent statistical limitations. However, our DTF analysis confirmed that scale-level bias remained modest despite item-level findings. Second, there is no consensus on thresholds for clinically meaningful DIF.^47^ We adopted sWABC < 0.10 as negligible based on established guidelines,^39,40^ but context-specific benchmarks for HRQOL measures in stroke populations remain to be established. Researchers must balance statistical detection against practical importance when interpreting DIF findings. Third, the AcT trial did not formally assess cognitive impairment, which could affect the reliability of self-reported HRQOL. While patients with severe pre-morbid disability were excluded, milder cognitive deficits common after stroke may have influenced responses. The impact of post-stroke cognitive impairment on EQ-5D-5L measurement properties deserves dedicated investigation. Fourth, the EQ-5D-5L is a generic HRQOL measure that may not capture stroke-specific concerns such as communication difficulties, fatigue, or emotional lability. DIF patterns in stroke-specific instruments warrant a separate study. Finally, while the age threshold of 80 years was consistent with prior AcT analyses, it was somewhat arbitrary. Sensitivity analyses with alternative cutoffs could provide additional insights, though our primary conclusions would likely remain unchanged given the modest magnitude of observed effects.

### Conclusions

In conclusion, the EQ-5D-5L demonstrates reasonable measurement invariance across age, sex, and treatment groups in acute ischemic stroke patients. Although statistically significant age-related DIF was detected in physical functioning items, effect sizes were modest and had minimal impact on individual-level HRQOL scores. The absence of meaningful DIF across sex and treatment groups supports the validity of EQ-5D-5L comparisons in stroke trials. These findings provide reassurance that the EQ-5D-5L can be used for HRQOL assessment and health economic evaluations in stroke populations without requiring group-specific adjustments.

## Data Availability

The datasets used and/or analyzed during the current study are available from the corresponding author upon reasonable request and with appropriate permission from the AcT trial team.

## Source of Funding

This study was supported by the Canadian Institute for Health Research, the Heart and Stroke Foundation of Alberta/University of Calgary, and the Alberta SPOR SUPPORT Unit.

## Disclosures

Dr. Menon reports stock in Circle NVI and patents for systems of triage in acute stroke. Dr. Sajobi reports consulting fees received from Circle NVI and employment at the University of Calgary. Dr Hill reports employment with the University of Calgary; consulting fees from Brainsgate Ltd; grants/contracts from Biogen, Inc, Boehringer Ingelheim, Canadian Institutes of Health Research, Medtronic, MicroVention, Inc, and NoNO, Inc; being part of the endpoint review committee at Merck; stock in Circle NVI; and patents for systems of triage in acute stroke. The other authors report no conflicts.

## Supplemental Material

**Table S1:** Response frequency distributions for EQ-5D-5L items (N = 1,264). **Table S2:** Local dependence assessment using Chen and Thissen’s G² statistic. **Table S3:** Item-level differential item functioning results for sex and treatment groups. **Figure S1:** Standardized residual correlations among EQ-5D-5L items.

## Nonstandard abbreviations and acronyms

AcT: Alteplase Compared to Tenecteplase
DIF: differential item functioning
DTF: differential test functioning
EQ-5D-5L: five-level EuroQOL questionnaire
GRM: graded response model
HRQOL: health-related quality of life
IRT: item response theory
mRS: modified Rankin Scale
NIHSS: National Institutes of Health Stroke Scale
PROMs: patient-reported outcome measures
sWABC: signed weighted area between curves
sDTF: signed differential test functioning
uDTF: unsigned differential test functioning

## Notes

### Clinical Trial

NCT03889249

### Author Declarations

The trial was regulated by Health Canada (Clinical Trials Application number 231509) and by research ethics boards at participating centres. The datasets used in your study had been de-identified prior to use in this study.

